# Improving Estimation of Intervention Impact on Antibody Titer Increase in the Presence of Missing Values: A Proposed Method for Addressing Limit of Detection Issues

**DOI:** 10.1101/2022.08.25.22279230

**Authors:** Yang Ge, W. Zane Billings, Amanda L. Skarlupka, Wannan Cao, Kevin K. Dobbin, Ted M. Ross, Andreas Handel, Ye Shen

## Abstract

In many laboratory assays that measure immunological quantities, a portion of the measured values fall below a limit of detection (LOD). This is also the case for the hemagglutination inhibition assay (HAI), a common method used to quantify antibodies in influenza research. The conventional approach is to treat values below the LOD as either equal to the LOD or LOD/2, which can introduce potential biases. These biases can become more pronounced when calculating compound measures such as the difference between post-vaccination and pre-vaccination antibody titers (titer increase). To address this issue, we conducted simulations using LOD measurements with LOD/2 values as the standard imputation. We then developed a new method to adjust coefficient estimates that account for the censored nature of measurements below the LOD. Applying this new method to data from an influenza vaccine cohort study, we compared the impact of vaccine dose on the titer increase of HAI.

**Author Summary:** Analysis of measurements obtained from widely used antibody assays frequently overlooks the underlying data structure, leading to potential biases in the results. To address this issue, we have developed a method that effectively reduces these biases.

## Introduction

The approval process for a new vaccine is often time-consuming.[1] Before undertaking costly and lengthy human trials, many vaccine candidates are compared using immunological markers. These markers serve as predictive indicators of vaccine efficacy (correlates of protection), enabling the allocation of resources to the most promising candidates.[2,3]

Antibodies are widely recognized as the most common correlates of protection. [4] Consequently, the change in antibody measurements before and after vaccination, also known as titer increase, is as an important indicator in various vaccine studies.[5–8] Antibody measurements, and many other immunological assays, produce discrete values in the form of dilution levels. These assays often have limits of detection (LOD) or quantification (LOQ), placing limits on the range of sample values that can be reliably measured. Specifically, values below an LOQ cannot be measured at the same precision as values above the threshold, while values below an LOD cannot be assigned a value by the assay. [9–11] Although many assays also have an upper LOD/LOQ, measures at the upper limit rarely occur in practice.

In studies of hemagglutination inhibition (HAI) assays for influenza antibodies, most studies treat the measured values as precise and impute all measurements below the LOD with either the LOD value or LOD/2 (standard imputation).[12] These values are then used for statistical analyses, which can compound the bias induced by constant imputation. For example, titer increase, defined as the ratio of post-vaccination titer to pre-vaccination titer, is often used in influenza vaccine studies to measure change from baseline, and can be compared to analyze the relative effects of interventions (e.g. placebo vs. vaccine, or vaccine A vs. vaccine B).

Researchers conduct statistical analyses to compare outcomes between study groups, with a particular focus on controlling for covariates known to influence the outcome, such as age. A specific example of this is the implementation of a linear regression model to assess the ratio of post-to pre-vaccination titers for two different types of vaccines (vac), while also controlling factors of age (age) and BMI (bmi). The equation would be given by log(𝑡𝑖𝑡𝑒𝑟_𝑝𝑜𝑠𝑡_/𝑡𝑖𝑡𝑒𝑟_𝑝𝑟𝑒_) = log(𝑡𝑖𝑡𝑒𝑟 𝑖𝑛𝑐𝑟𝑒𝑎𝑠𝑒) = 𝛽_0_ + 𝛽_1_ ∗ 𝑣𝑎𝑐 + 𝛽_2_ ∗ 𝑎𝑔𝑒 + 𝛽_3_ ∗ 𝑏𝑚𝑖 and the coefficient 𝛽_1_provides an estimate of the different impact of vaccines being compared. However, the standard LOD imputation introduces bias in the titer values leading to biased results of the model coefficients.[13–16]

Previous literature on data with LODs is mostly theoretical, and does not include examples with real data. [15,17,18] In this study, we used simulations to explore potential biases induced by the LOD, and propose and approach for adjustment to mitigate these issues. We also applied our method to analysis of HAI titers from a human influenza vaccine cohort study. [19]

## Methods

### Human cohort and simulated data

We used data from prior years of an ongoing vaccination study, which has been described in previous publications.[7,8,10] Briefly, from 2014–2018, the study recruited volunteers who had not yet received an influenza vaccine for the current season. Individuals under age 65 were given the standard dose (SD) FluZone vaccine (Sanofi Pasteur), while individuals aged 65 or older were given the choice between the SD vaccine or the recommended high dose (HD) FluZone vaccine. We limited our analyses to those individuals 65 or older, in order to accurately compared the two vaccines. Investigators collected samples before administering vaccinations and obtained follow-up samples with a target date of 21-28 days after the vaccinations. The serum samples were then analyzed using HAI assays as described in a prior study. [10]

### HAI titer data

The main outcome measure we considered was the titer increase after vaccination, calculated as the logarithm of the ratio between post-vaccination and pre-vaccination titers:

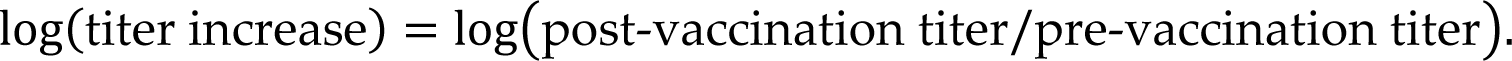

We use the notation log(𝑥) to represent the natural logarithm. In our dataset, the lowest dilution limit was set at 1:10, and subsequent dilutions were made in 2-fold increments (1:20, 1:40, and so on) up to the highest dilution of 1:20480. Measurements are generally reported as the reciprocal titer, such as 10, 20, 40, and so on. Any measurement that fell below the limit of detection (LOD) at the lowest reciprocal titer (10) was recoded as 5 using the standard imputation method. Since there were no values at the upper limit of the assay in our observed data (the highest titer was 5120), and these values are generally rare in similar studies, we only considered the lower LOD in our study.

### Simulated data

In our simulations, we generated data that resembles actual HAI assay data. We also computed the titer increase and used the classic linear regression model for the estimation of variables’ coefficients. In this simulation, we chose one intervention and one covariate motivated by the real data described above. The intervention was a binary variable (HD or SD influenza vaccine), which we represented as an indicator variable where 0 indicates SD and 1 indicates HD. We chose to simulate age as an integer-valued covariate. Thus, our regression model was log(𝑡𝑖𝑡𝑒𝑟 𝑖𝑛𝑐𝑟𝑒𝑎𝑠𝑒) = 𝛽_0_ + 𝛽_𝑣_ ∗ 𝑣𝑎𝑐𝑐𝑖𝑛𝑒 + 𝛽_𝑎_ ∗ 𝑎𝑔𝑒.

We generated pre-vaccination antibody titer values with log-normal distributions (which we call the raw titers). The means of these distributions were chosen to produce five percentages (10%, 30%, 50%, 70%, 90%) of values that would be below the LOD (10). Next, we discretized all values by rounding them down to the nearest dilution level. For instance, any value between 20 and 40 was coded as 20 (referred to as Raw+bin). Finally, we assigned a value of 5 to any measurement below the assay’s limit of detection of 10 (referred to as Raw+bin+lod5).

We then calculated post-vaccination titers for each of the three types of pre-titers (raw, Raw+bin, or Raw+bin+lod5). We then specified the expected titer increase for each of the vaccine groups. After setting the mean of the pre-vaccination titer distribution and the expected titer increase value, we calculated the necessary mean for the post-vaccination titer distribution and use these values in our simulations.

As HD is expected to induce a higher titer increase than SD,[20] we assumed the log(titer increase) for HD and SD to be log(12) and log(6) , respectively, as a representative example. So, the difference of titer increase between HD and SD (true coefficient value) was log(12) − 𝑙𝑜𝑔(6) = 0.7. Similarly, as an example, we chose the coefficient value of age to be -0.03 per 1 year older, considering that the older population may have a lower titer increase.[10,21] Sensitivity analyses for these parameters are provided in the supplementary materials.

The true values of HAI titer values that we observe to be below the LOD are unknown, so we conducted additional simulations to better explore potential bias. More specifically, we performed 500 simulations for each of the three types of simulated datasets (i.e., raw, Raw+bin, or Raw+bin+lod5), with each simulation consisting of 1000 samples. Next, we separately fit linear models to the three types of simulated datasets using both the standard imputation method and the method we proposed (described in the next section). Finally, we compared the coefficient estimates obtained from these two methods.

### Adjustment for Bi-Censoring (ABC) method

Often, pre- and post-vaccination antibody titers are measured for different scenarios (e.g., two types of vaccines) and the ratio (log difference) of the titers, called the titer increase, is compared using classical linear regression models or other standard statistical methods. However, censored values are common in HAI assays due to the limit of detection (LOD). This bi-censoring phenomenon affects both pre-vaccination and post-vaccination HAI titers. Without proper adjustments, models may treat values below the LOD as fixed and accurate, leading to biased coefficient estimates.

We found the titer increase had bi-censoring issues, where the values of titer increase may be treated as interval censors.[13] Therefore, to address this, we developed an adjustment method called *Adjustment for Bi-Censoring* (ABC). HAI antibody titers that show up as negative in the assay, indicating values below the LOD, can span a range from just below the LOD to zero. Due to maternal immunity and cross-reactivity, it is rare for individuals to have completely zero immunity to a specific strain of influenza virus.[22,23] The low bound of the range is more likely to be a low but non-zero level.[24] Therefore, we assume those values below LOD could be any value between 1:10^−5^ to 1:10. The threshold of 10^−5^ was chosen so that the range covered the majority part of the log-normal distribution for those values below LOD. Since the pre-vaccination and post-vaccination HAI titer values were left-censored, and values below the LOD were within a range, the estimation of coefficients for the titer increase can be approached as an interval censoring problem. The log-likelihood function can be maximized to obtain the coefficient estimates:

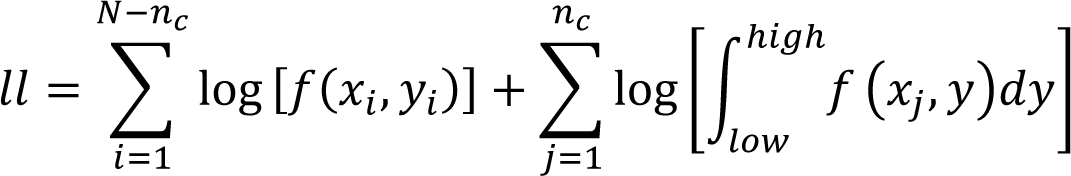

where x is the intervention; y is the titer increase; N is the total sample size; 𝑛_𝑐_ are values below LODs; log[𝑓(𝑥_𝑖_, 𝑦_𝑖_)] is the log of the density of normal distribution with mean ℎ𝑖𝑔ℎ equal to 𝜇 , variance equal to 1; 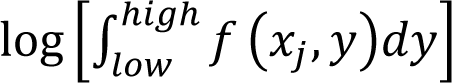 is the cumulative probability function between the high and low bound of the same normal distribution.

To compare the proposed ABC method with the standard imputation method (benchmark), we calculated the squared error loss (SEL). The SEL is the sum of the variance and squared bias, which is derived from the difference between the coefficient assumption 𝛽 and the estimated coefficient ^𝛽^^ using the squared error loss (bias-variance decomposition).[25,26]

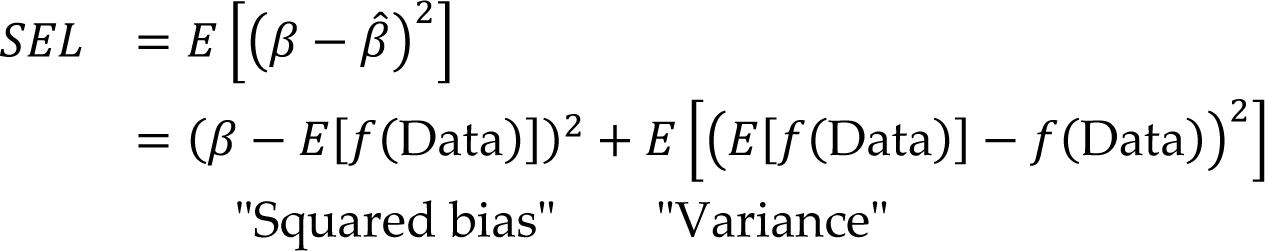

### Sensitivity analyses

We conducted sensitivity analyses by varying the sample size of simulated data (N = 100, 500, 1000, 1500), and two different coefficient assumptions (log(24/6), log(6.5/6)) for the difference of titer increase between two vaccines, as well as the impact of age (Supplementary material).

### Model Implementation

All models and analyses were implemented in R.[27] To maximize the log-likelihood, we used the Newton-Raphson method and the maxLik package.[28,29] All the code and data required to reproduce the results are provided in the supplementary material.

## Results

We generated a series of simulated datasets with LOD percentages at five levels (10%, 30%, 50%, 70%, 90%), and for each level, we repeated the simulation 500 times. Figure 1-A visualizes a dataset at one LOD level. The simulated pre- and post-vaccine titers demonstrated the effects of binning and LOD imputation on the central tendency and dispersion of the data. Figure 1-B to D show the simulated data in all three scenarios. The first scenario (Figure 1-B) is the raw data with continuous values; the second scenario (Figure 1-C) is the raw+bin data with discrete integers; the last scenario (Figure 1-D) is the raw+bin+lod5 data where titers below LOD (1:10) were recoded as 1:5. For both HD and SD groups, the raw+bin+lod5 data had a higher mean and small variance of titers than the other two scenarios. Figure 1-E represents the titer increase calculated as the ratio between post-vaccine and pre-vaccine titers. The distribution of titer increase changed in raw+bin+lod5 data of both HD and SD groups because of the standard imputation method.

**Figure 1:**
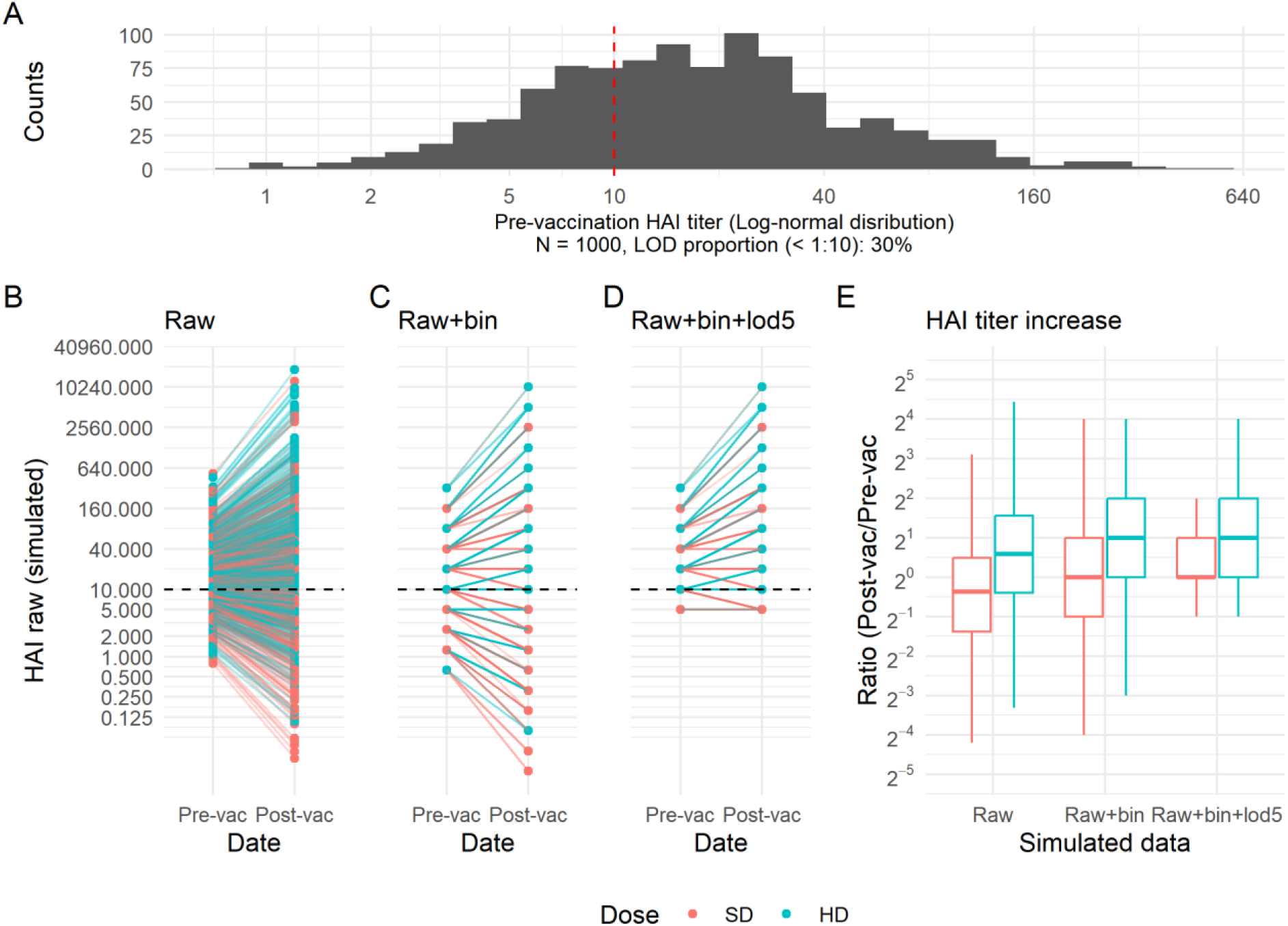
Simulated HAI titer and LOD. A, example of a simulated log-normal distributions with a total sample size equal to 1000 with about 30% values below LOD; B, the simulated raw data; C, the simulated raw+bin data (binning raw data into discrete integers); D, the simulated raw+bin+lod5 data, introducing the LOD issue (<1:10 coded as 1:5) into the simulated raw+bin data; E, HAI titer increase for both HD and SD vaccines using the three versions of simulated data.

In our assumptions, we set the parameter for the difference in titer increase between the two vaccines to 0.7 (𝑙𝑜𝑔(𝐻𝐷_𝑡𝑖𝑡𝑒𝑟 𝑖𝑛𝑐𝑟𝑒𝑎𝑠𝑒_/𝑆𝐷_𝑡𝑖𝑡𝑒𝑟 𝑖𝑛𝑐𝑟𝑒𝑎𝑠𝑒_) = 𝑙𝑜𝑔(12/6)), reflecting a higher titer increase for the HD vaccine compared to the SD vaccine. Additionally, we assigned a parameter value of -0.03 for age (per 1-year unit increase), taking into account the lower immune response typically observed in the elderly population. (sensitivity analyses are provided in supplementary material). By fitting a classic linear regression model to the three versions of simulated data, we observed that the linear regression models using the raw and raw+bin data produced results that were close to the assumed values (Figure 2 and 3, black and yellow bars). However, when the data had LOD issues (raw+bin+lod5 data), the standard imputation method resulted in biased coefficient estimations for the model. (Figure 2 and 3, blue bars). The magnitude of bias increased as the proportion of data below the LOD increased.

**Figure 2:**
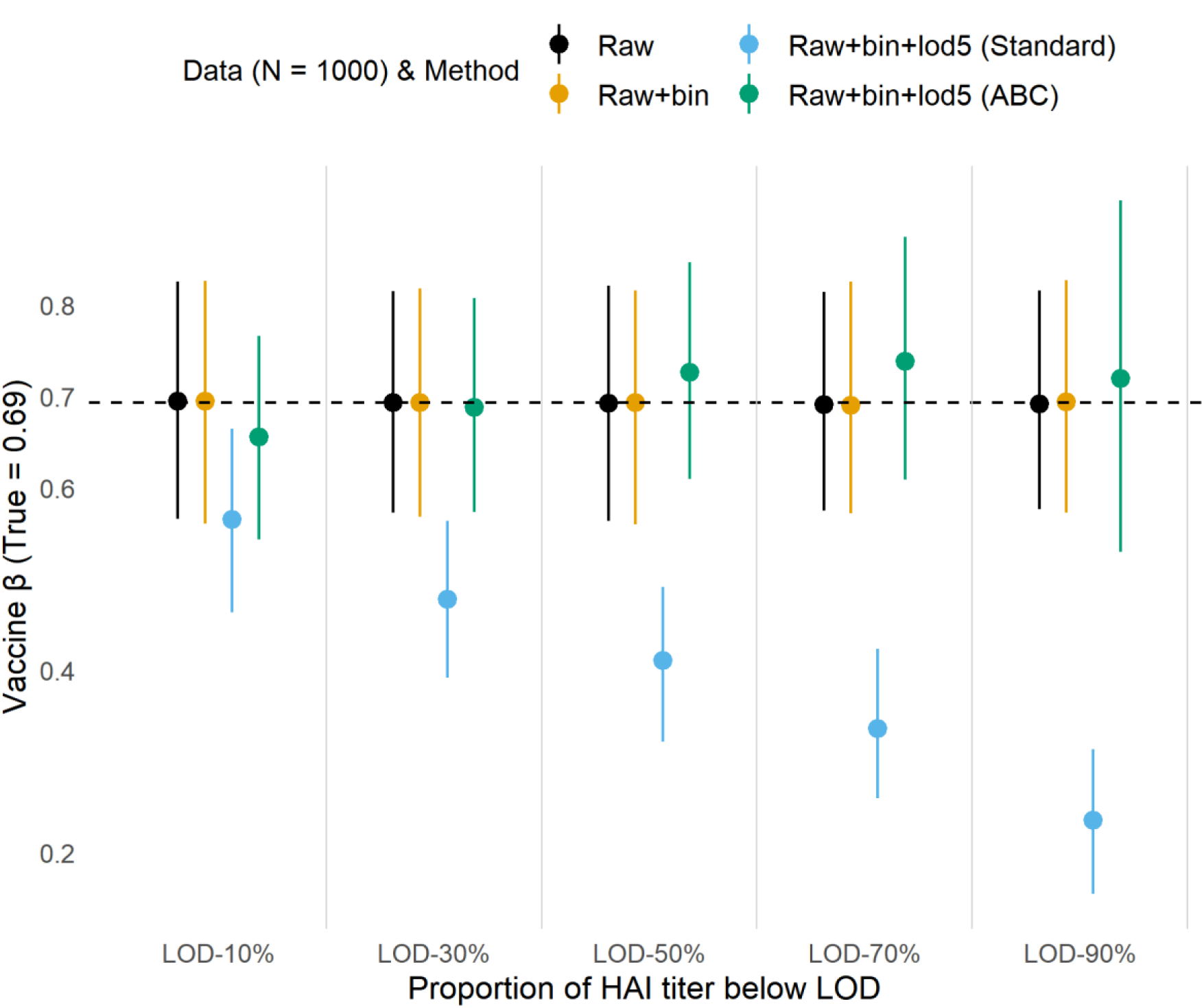
Coefficient estimates of the difference of titer increase (HD vs. SD vaccine). We generated 500 datasets, each with a sample size (N) of 1000, to estimate the mean coefficient and its 95% confidence interval. We used three types of data in our analysis: raw data, which represents continuous titer values; raw + bin data, which represents discrete titer values; and raw + bin + lod5 data, which represents discrete titer values with LOD standard imputation. The Adjustment for Bi-Censoring (ABC) method was applied to the raw + bin + lod5 data for adjustment.

**Figure 3:**
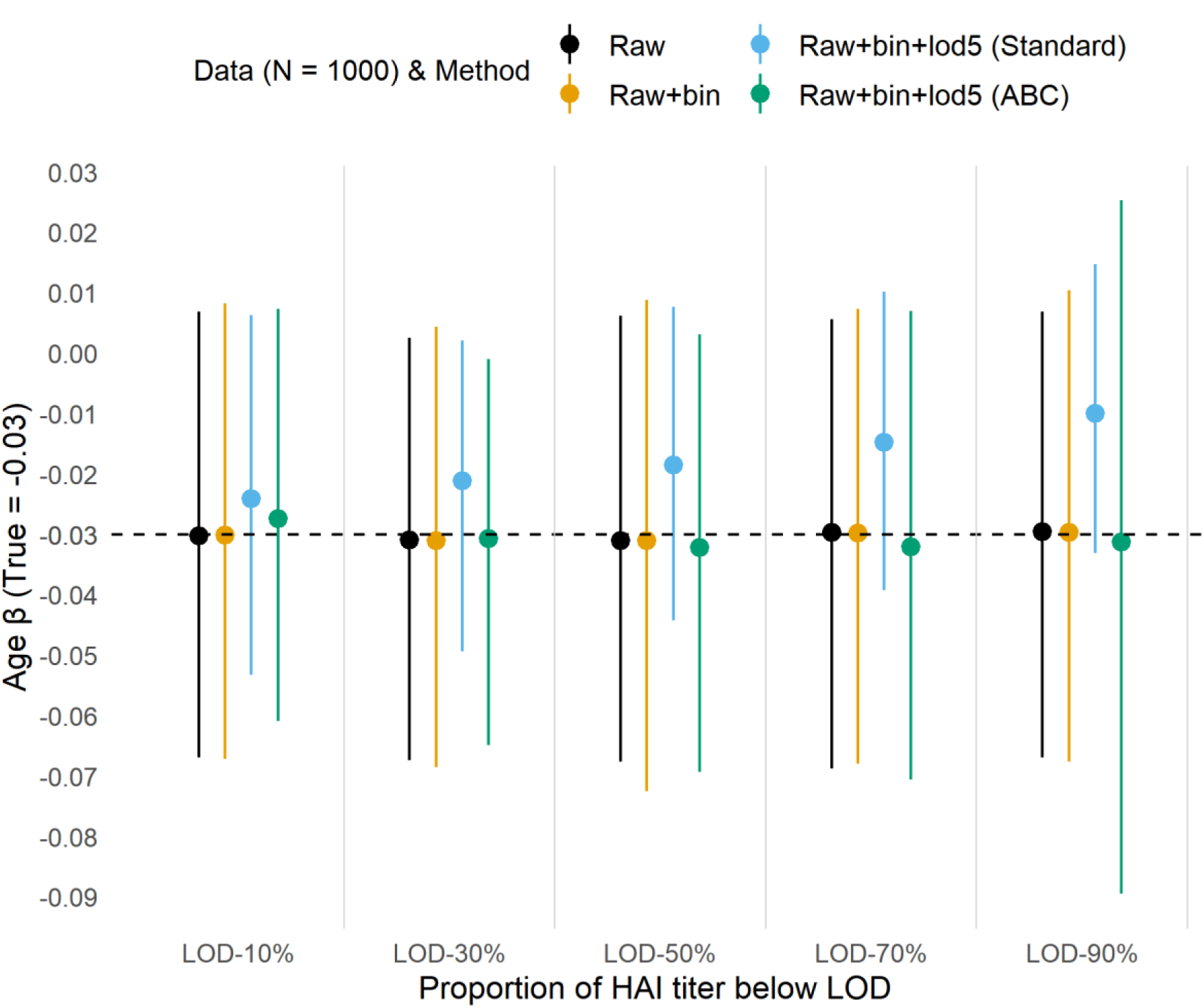
Coefficient estimates of the age (every 1-year increase). There were 500 datasets, each had N = 1000 as the sample size, for coefficient mean and 95% confidence interval estimation. We used three types of data in our analysis: raw data, which represents continuous titer values; raw + bin data, which represents discrete titer values; and raw + bin + lod5 data, which represents discrete titer values with LOD standard imputation. The Adjustment for Bi-Censoring (ABC) method was applied to the raw + bin + lod5 data for adjustment.

### Bias–variance trade-off

The ABC method yielded results with reduced bias compared to the standard imputation method. However, it was accompanied by higher variance, which can be attributed to the increased proportion of values below the LOD. Thus, we further explored the Bias– variance trade-off (Figure 4). As the sample size increased, the sum of squared bias and variance decreased. However, it increased as the proportion of values below the LOD (limit of detection) increased. The ABC method had less bias-variance score than the standard imputation method in most scenarios, except when the sample size was small. This is because the standard imputation method ignored the uncertainty due to LOD.

**Figure 4:**
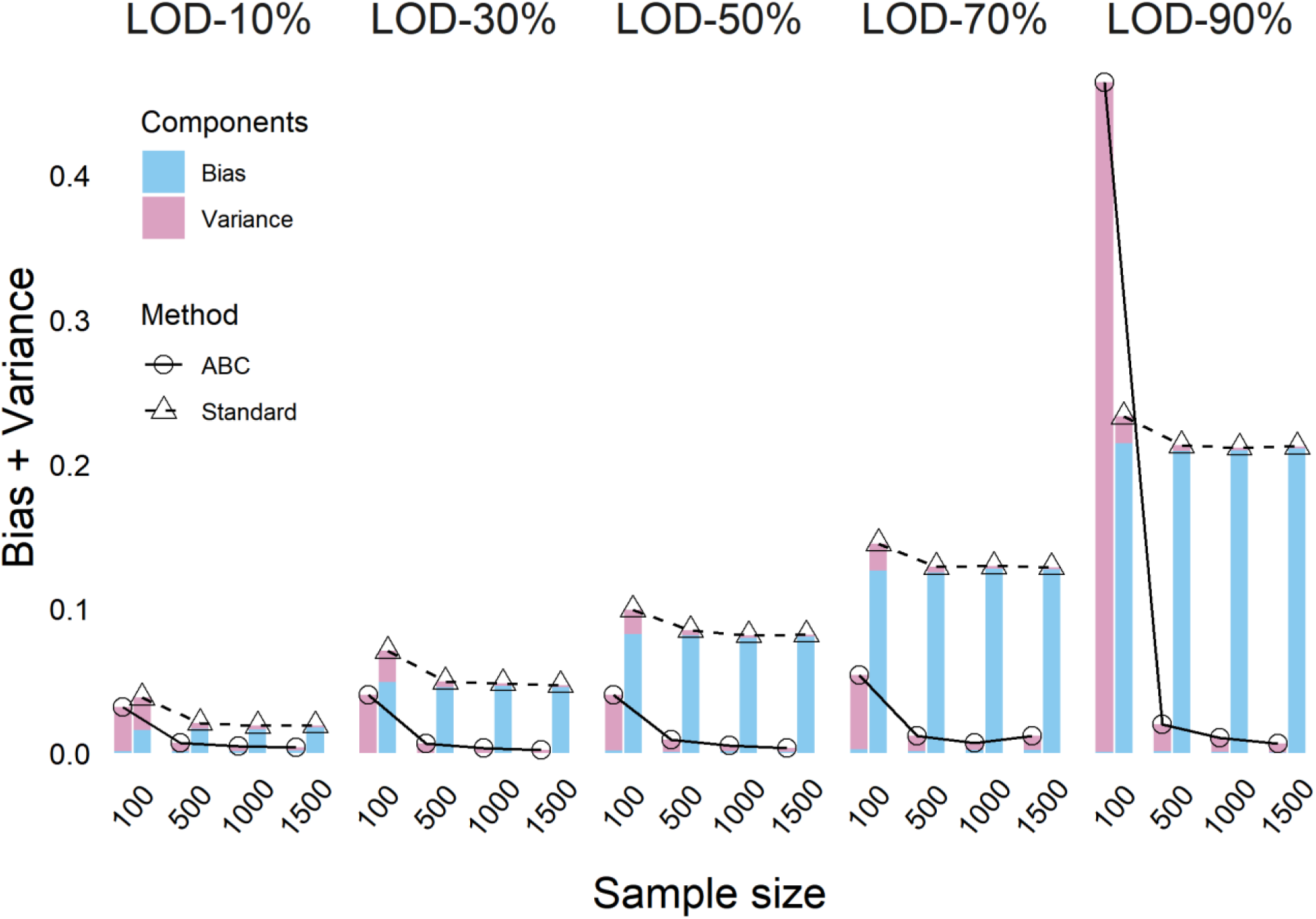
Bias–variance trade-off score with different LOD and sample size settings. To calculate the score, we simulated 500 datasets with multiple scenarios, including different sample sizes (N = 100, 500, 1000, 1500) and proportions of values below LOD (10%, 30%, 50%, 70%, 90%).

### Apply the ABC method to the human cohort data

We also explored the ABC and the standard imputation method with the real human cohort data to compare their results. For vaccine homologous (HAI titer against the same strain included in the vaccine) comparisons, the ABC and standard method provided different results for several strains (Table 1). For example of H1N1-California-2009 and H1N1-Michigan-2015 strains, the estimation of coefficients changed from significant in the standard imputation method to non-significant with the ABC method. For vaccine heterologous (HAI titer against a strain not included in the vaccine) comparisons, these two methods provided similar results, but the ABC method had wider confidence intervals (Table 2). For example of H1N1-Brisbane-2007 strain, the proportion of pre-vaccination titer below the LOD was 100%. Therefore, the ABC method provided a wide interval. For the H3N2-Hong Kong-2014 strain, the result was significant with the ABC method, but was non-significant with the standard imputation.

**Table 1:** Comparison of model estimations on selected homologous responses

| Vaccine strain | LOD | Standard method | ABC method |
| --- | --- | --- | --- |
| H1N1-California-2009 | 26.2% | 0.31, 95%CI: 0.11 to 0.51 | 0.23, 95%CI: -0.02 to 0.48 |
| H1N1-Michigan-2015 | 7% | 0.45, 95%CI: 0.01 to 0.89 | 0.44, 95%CI: -0.13 to 1.01 |

**Table 2:**
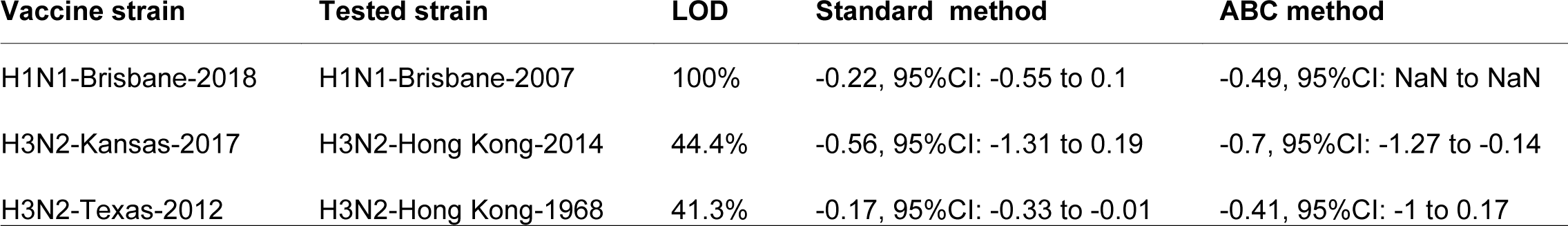
Comparison of model estimations on selected heterologous responses

| Vaccine strain | Tested strain | LOD | Standard method | ABC method |
| --- | --- | --- | --- | --- |
| H1N1-Brisbane-2018 | H1N1-Brisbane-2007 | 100% | -0.22, 95%CI: -0.55 to 0.1 | -0.49, 95%CI: NaN to NaN |
| H3N2-Kansas-2017 | H3N2-Hong Kong-2014 | 44.4% | -0.56, 95%CI: -1.31 to 0.19 | -0.7, 95%CI: -1.27 to -0.14 |
| H3N2-Texas-2012 | H3N2-Hong Kong-1968 | 41.3% | -0.17, 95%CI: -0.33 to -0.01 | -0.41, 95%CI: -1 to 0.17 |

### Sensitivity analyses

We conducted sensitivity analyses by varying the sample size of the simulated data, and changing the assumptions of coefficients (titer increase and age). Across all scenarios, we consistently observed less biased estimates with the ABC method. Detailed results can be found in the supplementary material.

## Discussion

Our ABC method addresses the issue of measurements below the LOD, which is applicable beyond influenza HAI titers. Most antibody assays have an LOD, and observations below this limit are common in practice. [5–11] In a human influenza vaccine cohort, we observed relatively low proportions of LOD measurements in homologous responses. However, in heterologous responses, the proportions of LOD measurements were significantly higher. Our findings demonstrate that failing to properly account for LOD measurements can lead to biased results.

We proposed the ABC method that treated values below the LOD as censored, and incorporates this information into the estimated likelihood. Our ABC method reduces the bias of coefficient point estimates, while incorporating the additional uncertainty induced by censored values into confidence intervals, providing a more honest assessment of model error. Comparing ABC to the standard constant imputation method, as the proportion of LOD increased, the bias of coefficient estimations with the standard imputation method increased, but the estimations with the ABC method did not. In addition, we aimed to propose an adjustment that had low bias and variance.[25,26] While the standard imputation method can often yield estimates with smaller variance, the smaller variance is dishonest, as it treats all values below the LOD as if they were perfectly observed at the same constant level, reducing uncertainty in model estimates. The variance of coefficients can depend on 1) the proportion of LOD in the data, 2) the total sample size of the data, and 3) the effect size of the intervention. However, as the sample size increases, the variance of the two methods was similar even for data with many values below the LOD.

While the ABC method can generate unbiased coefficient estimates, our method has several model limitations. First, sensitivity analyses revealed that when the proportion of values below LOD was high, the ABC method could result in wide confidence intervals, leading to inconclusive results. Second, the current version of the ABC method does not account for hierarchical structured data, which is an area we aim to address in future research. Finally, our method requires the use of standard numerical optimization procedures, which are slower and more computationally intensive than the fast methods used for standard linear models.

Our findings may benefit antibody comparisons between vaccine candidates. We recommend the use of our ABC method, which addresses the concern of LOD, as an alternative to the standard method.

## Data Availability

All data of this study are provided in the Supplementary Materials.

